# Is Google Trends a useful tool for tracking mental and social distress during a public health emergency? A time-series analysis

**DOI:** 10.1101/2021.02.18.21251966

**Authors:** Duleeka Knipe, David Gunnell, Hannah Evans, Ann John, Daisy Fancourt

## Abstract

**Background:** Google Trends data are increasingly used by researchers as an indicator of population mental health, but few studies have investigated the validity of this approach.

**Methods:** Relative search volumes (RSV) for the topics depression, anxiety, self-harm, suicide, suicidal ideation, loneliness, and abuse were obtained from Google Trends. We used graphical and time-series approaches to compare daily trends in searches for these topics against population measures of these outcomes recorded using validated scales (PHQ-9; GAD-7; UCLA-3) in a weekly survey (n=∼70,000) of the impact COVID-19 on psychological and social experiences in the UK population (12/03/2020 to 21/08/ 2020).

**Results:** Self-reported levels of depression, anxiety, suicidal ideation, self-harm, loneliness and abuse decreased during the period studied. There was no evidence of an association between self-reported anxiety, self-harm, abuse and RSV on Google Trends. Trends in reported depression symptoms and suicidal ideation declined over the study period, whereas Google topic RSV increased (p=0.03 and p=0.04 respectively). There was some evidence that suicidal ideation searches preceded reported self-harm (p=0.05), but graphical evidence suggested this was an inverse association. However, there was statistical and graphical evidence that self-report and Google searches for loneliness (p<0.001) tracked one another.

**Limitations:** No age/sex breakdown of Google Trends data are available. Survey respondents were not representative of the UK population and no pre-pandemic data were available.

**Conclusion:** Google Trends data do not appear to be a useful indicator of changing levels of population mental health during a public health emergency, but may have some value as an indicator of loneliness.

## Background

Google Trends data are free and easily accessible, allowing near real-time tracking of Google search activity on a range of issues. In recent years, there has been growing interest in their usage in suicide research (Gunnell et al., 2015; Nuti et al., 2014) and other mental health research (Ayers et al., 2012; Bragazzi, 2013; Tefft, 2011; Yang et al., 2010). During the COVID-19 pandemic, the utilisation of this tool to track changing levels of population distress and public concerns has proliferated (Ayers et al., 2021; Hoerger et al., 2020; Knipe et al., 2020a; Rana, 2020; Sinyor et al., 2020). For example, a review of records identified in our Living Review of the impact of COVID-19 on suicidal behaviour identified 13 such studies up to 31 January 2021 (John et al., 2020). However, concerns have been raised about its usefulness as a mental health surveillance tool (Arora et al., 2019; Page et al., 2011; Tran et al., 2017).

Whilst the easy availability of Google search data makes it deceptively simple to access and analyse, there are a number of potential pitfalls. First, using Google search data as a measure of population level experiences relies on the assumption that most searching on mental health or suicide-related terms is carried out by individuals having such experiences or thoughts themselves. However, search activity may, for example, be triggered by curiosity following news reports of suicide deaths, particularly celebrity deaths. Furthermore, the sociodemographic characteristics of internet users – more often young people with access to the Internet – are likely to under-represent the elderly and those in digital poverty largely from socioeconomically deprived or rural backgrounds. Despite being a large dataset, its relevance for population-level tracking of experiences remains questionable. Finally, it is unclear to what extent individuals experiencing poor mental health or suicidal thoughts will necessarily turn to Google searching, and whether such searching is contemporaneous with their thoughts and experiences or lagged (e.g. occurring after several days of symptoms). These limitations all have the potential to seriously affect the reliability of using Google search activity within mental health research.

To date, few studies have investigated associations of Google search activity for mental health issues with outcomes other than suicide. A recent cross-national study (n=202 countries) reported cross-sectional associations between the United Nations Happiness Index and national relative search volumes for anxiety (r=0.39 p<0.001) in 2017) and happiness (r=0.17 p<0.05 in 2017) (Banerjee, 2018). However, the authors did not look at changes over time, and the validity of cross-national comparisons of Google search volumes is questionable given the way Google normalises its data. Much of the literature has focused on the association between Google searches for terms indicating suicide risk (e.g. depression / suicide methods / suicide help) and changes in population suicide rates (Arora et al., 2019; Ayers et al., 2012; Ayers et al., 2021; Barros et al., 2019; Gunnell et al., 2015; Hoerger et al., 2020; Knipe et al., 2020a; McCarthy, 2010; Misiak et al., 2020; Nuti et al., 2014; Rana, 2020; Sinyor et al., 2020; Sueki, 2011; Tran et al., 2017; Yang et al., 2010). Findings from these studies are mixed in the associations they report. A recent analysis of Google Trends and suicide data over a 10-12 year period for Austria, Germany, Switzerland and the USA using multiple search terms found little evidence of consistent associations across different countries and recommended caution in the use Google Trends data for forecasting suicide trends (Tran et al., 2017). In contrast, in an analysis of Google Trends, unemployment and suicide data for Ireland, Barros and colleagues concluded that: “the combination of societal data and online behaviour provide a good indication of societal [suicide] risks” (Barros et al., 2019). In an analysis of US suicide and Google trends data for 2004-2007, McCarthy reported that whereas Google searches for the term “suicide” were inversely associated with suicide and self-injury rates in the overall population, the opposite association was seen in an analysis of searching for “teen suicide” vs. suicide and self-injury rates in young people; the analysis was restricted to data for four years only and so power was limited (McCarthy, 2010). In keeping with the possible age-sensitivity of findings, and the fact that younger people are greater users of the Internet than older individuals, an analysis of Google Trend and suicide data for England and Wales (2004-2013) found that whereas the correlation with overall population suicide rates was weak (r=0.16), it was highest in 25-34 year olds (r=0.85), although low in 15-24 year olds (r=0.29) (Arora et al., 2019). However, such studies have focused on Google trends and suicidal acts. They do not tell us about whether Google search behaviours are related to experiences or feelings that individuals might be having that could reflect broader mental health experiences or act as a precursor to suicide attempts. Indeed, we are aware of no previous studies investigating associations between Google search activity and changing levels of other indicators of mental health (e.g. depression, anxiety, self-harm) and social distress (e.g. loneliness, physical and psychological abuse).

This study investigates the relationship between UK Google Trends data and self-reported mental health and social distress indicators in a large cohort study involving repeated surveys of over 70,000 adults across the UK. Specifically, we focused on the period from 21 March 2020 up to 21 August 2020, which coincided with the start of the UK’s first lockdown due to the COVID-19 pandemic. This specific context is pertinent for exploring the relationship between mental health and Google search behaviour given it covered a time where population trends in mental health changed markedly as a result of fluctuating concerns about COVID-19 and changes in population social and economic circumstances resulting from the public health measures to contain the spread of the virus (period of extreme lockdown restrictions 23 March – 17 May 2020). Therefore, this provided marked changes in population averages for mental health in in measures that are often relatively stable across the population (Fancourt et al., 2021).

## Methods

### Data

This study used data from the COVID-19 Social Study; a large panel study of the psychological and social experiences of adults (aged 18+) in the UK during the COVID-19 pandemic. The weekly online longitudinal survey started on the 21/03/2020 and has collected data from over 70,000 adults. Participants were asked to complete a survey once a week across a 24-week period. For details on the recruitment, sampling, retention and weighting of the sample is available in the study user guide: https://github.com/UCL-BSH/CSSUserGuide. For this analysis, all survey responses between 21/03/2020 and 21/08/2020 were analysed.

Age, sex, and socioeconomic information were collected as well as data on a range of mental and social distress measures (Iob et al., 2020). Depression was assessed using a modified version of the nine-item Patient Health Questionnaire (PHQ-9) (Kroenke et al., 2001), and anxiety was assessed using the modified version of the seven-item Generalised Anxiety Disorder Assessment (GAD-7) (Spitzer et al., 2006). The original PHQ-9 and GAD-7 questionnaires refer to a time period of the last two weeks. In this COVID-19 Social Study, the time period is the last week. Although PHQ-9 and GAD-7 are not diagnostic tools, they have both been used to assess depression and anxiety in the general population (Kroenke et al., 2001; Spitzer et al., 2006). We used item 9 in the PHQ-9 to assess population levels of suicidal/self-harm thoughts: “Over the last week, how often have you been bothered by thoughts that you would be better off dead, or of hurting yourself in some way?”. Self-harm was assessed by asking “Over the last week, how often have you been bothered by self-harming or deliberately hurting yourself”. Psychological and physical abuse (referred to as abuse) was measured by asking two questions “Over the last week, how often have you been bothered by being physically harmed or hurt by somebody else?” and “Over the last week, how often have you been bothered by being bullied, controlled, intimidated or psychologically hurt by someone else?”. All questions were asked on a four-point scale from “not at all” to “nearly every day”. Loneliness was measured using the three-item UCLA loneliness scale (UCLA-3) (Russell, 1996).

We used *Google Trends* data to track Google searches over time in the UK. Google Trends provides daily relative search volume (RSV) data for specific search terms and Google-defined topics. Topics are a group of related terms (defined by Google) that share the same concept in any language. Google does not provide information on the absolute numbers of searches; rather the RSV is first normalised by dividing each data point by the total searches for the specified time range and geographical area. The resulting number is then indexed, where 100 is the maximum search interest for the topic during that time in that location. Periods with very low search volumes are identified as zero activity.

The time period used for downloading data from Google spanned the period of strict COVID-19 pandemic-related lockdown measures in the UK followed by the easing of such measures over the summer (21/03/2020 - 48 hours before national restrictions were brought in – to 21/08/2020). This period was consistent with the time period for which we have self-reported data. RSVs for the topics identified in our previous analysis with corresponding self-reported data in the COVID-19 Social Study were downloaded (Knipe et al., 2020a). Daily RSVs for the topics depression, anxiety, self-harm, suicide, loneliness, and abuse were downloaded. In addition, we downloaded data on the topic of suicidal ideation as this was measured in the COVID-19 Social Study. All searches used Google topics (not terms – see above), included all Google query categories and included all web searches (i.e. includes image, news, Google shopping, and YouTube searches) in the UK. Previous analysis of Google Trends data has highlighted that slightly different RSVs are provided by Google for the same search (with the same parameters) on different days (Tran et al., 2017). We therefore downloaded data on seven different days using the parameters specified (detailed above) and created an averaged dataset. We took the average value for each search topic (7 topics) on each of the 154 datapoints, as it was recorded on each of the 7 separate days (working days between 14/01/2021 – 22/01/2021).

### Statistical analysis

All analyses were conducted using STATA 16 (StataCorp, 2019). We used previously recommended cut-offs for the validated scales (Kroenke et al., 2001; Spitzer et al., 2006) – a PHQ-9 or GAD-7 score of 10 or more was used to indicate moderate/severe depression or anxiety symptoms. For suicidal ideation, self-harm, and abuse, a response that indicated at least one occasion of these in the previous week was recorded as an experience of these thoughts/events. For each day of the study period (21/03/2020-21/08/2020) the proportion of responses indicating depression, anxiety, suicidal ideation, self-harm, and abuse were generated. The mean score of the UCLA-3 scale (loneliness measure) was calculated for each day.. The questions related to self-harm, and abuse were only collected from the 30/03/2020 onwards.

We provide graphical presentations of Google Trends topic RSVs and self-reported measures of mental and social distress. Our analysis aimed to estimate the temporal association of one time series on another. As we did not have data on suicide deaths and attempts during this period, we investigated associations between Google searches for suicide with self-reported self-harm and suicidal ideation. Given the low likelihood that an individual’s Google searching for a mental or social distress term will result in them developing or experiencing distress, we assumed that the development of symptoms or experience of abuse would predate Google search behaviour and not occur simultaneously. The only exceptions to this might be Google searches for terms related to suicidal thoughts, self-harm and suicide which may precede self-reported self-harm. We used vector autoregressive (VAR) to test whether there was evidence that one time series temporally preceded another. These models account for autocorrelation, and allows for lags in effect (Becketti, 2013). We observed a day of the week effect in Google searches for topics, and therefore added dummy variable in for day of the week as an exogenous variable to account for this. We estimated two-variable VARs each using the seven self-reported mental/social distress data with the corresponding Google Trends time series. For each VAR we needed to select the number of lags to estimate our models. We did this by selecting the best fitting model by testing out a range of lag lengths by using the *varsoc* command in STATA and used the Akaike’s information criterion to select the number of lags to estimate the VAR models (see Table 1). VAR models were fitted using the *var* command. All models were checked for stationarity. We used the Granger causality test to assess whether the self-reported time series predict Google Trend values for the corresponding mental/social distress topics. In addition, given the possibility that Google searches for topics related to suicidal ideation might precede self-harming behaviour we also tested for this using a Granger causality test.

**Table 1.** Granger causality test results for the association between self-reported and Google searching time trends data

|  | No. of lags<br>used in VAR<br>models<sup>a</sup> | Granger causality test p-values |  |
| --- | --- | --- | --- |
|  |  | Self-reported data precedes<br>Google searching | Google searching precedes<br>self-reported data |
| **Mental distress** |  |  |  |
| Depression | 2 | 0.03 | - |
| Anxiety | 3 | 0.62 | - |
| Suicide ideation<sup>b</sup> |  |  |  |
| Self-harm | 2 | 0.12 | 0.05 |
| Suicidal<br>ideation | 3 | 0.74 | - |
| Self harm<sup>b</sup> |  |  |  |
| Self-harm | 2 | 0.28 | 0.43 |
| Suicidal<br>ideation | 3 | 0.97 | - |
| Suicide<sup>b</sup> |  |  |  |
| Self-harm | 2 | 0.26 | 0.67 |
| Suicidal<br>ideation | 4 | 0.04 | - |
| **Social distress** |  |  |  |
| Loneliness | 2 | <0.001 | - |
| Abuse | 3 | 0.11 | - |

<sup>a</sup>VAR – Vector autoregressive. The number of lags is the number of days between one time trend (e.g. self-reported) and the other (e.g. Google searches).
<sup>b</sup>Google search topics were compared to self-reported self-harm and suicidal ideation time trends data

Given the age patterning of mental/social distress and internet use, we also provide graphical presentations of Google Trend topic RSVs and self-reported measures of mental and social distress stratified by age group (18-29; 30-59; 60+).

### Ethics

The UCL Social Study survey was approved by the UCL Research Ethics Committee and all participants gave written informed consent.

## Results

72,046 individuals responded to the UCL Social Study survey and provided data on at least one social or mental distress measure during the study period. The majority of respondents were female (75%), with an average age of 49 years (SD 14.9) and were university graduates (67%). This analysis included data from 675,651 surveys (mean 4423 per day within the study period, SD 1271) with responses to at least one mental or social distress measure.

Temporal trends in Google search activity and self-reported measures of mental distress over the study period are shown in Figure 1. Rates of self-reported depression, anxiety, suicidal ideation, and self-harm declined by 5-44% over the 5-month study period. However, relative Google searches for these topics remained fairly stable, with the exception of depression and suicidal ideation, which appeared to increase during the early part of the study period. Self-reported levels of loneliness and abuse appeared to drop during the pandemic, Google Trend data appeared to follow the trends for loneliness but not abuse (Figure 2).

**Figure 1.**
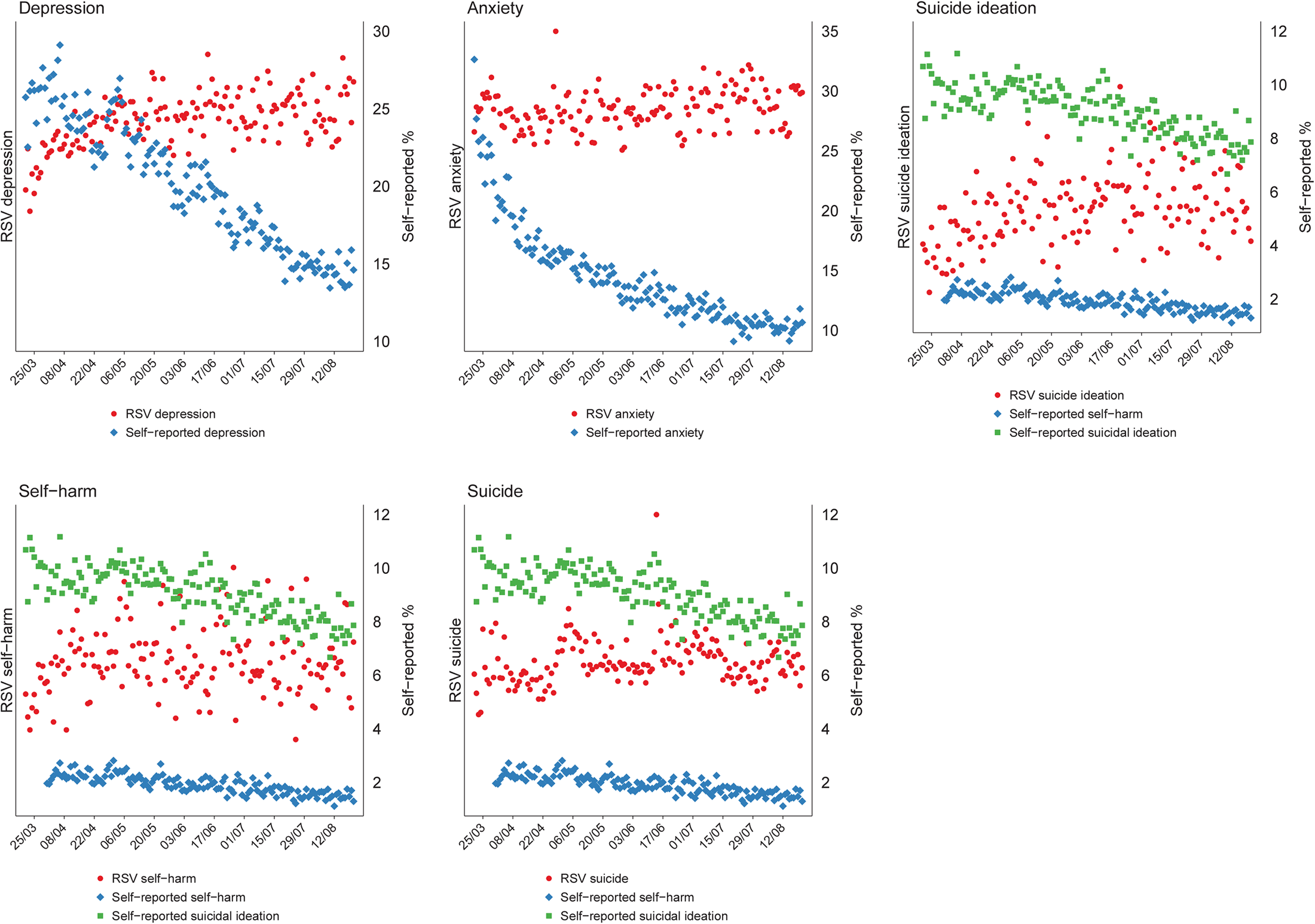
Self-reported and Google Trend search data for mental distress topics in the UK

**Figure 2.**
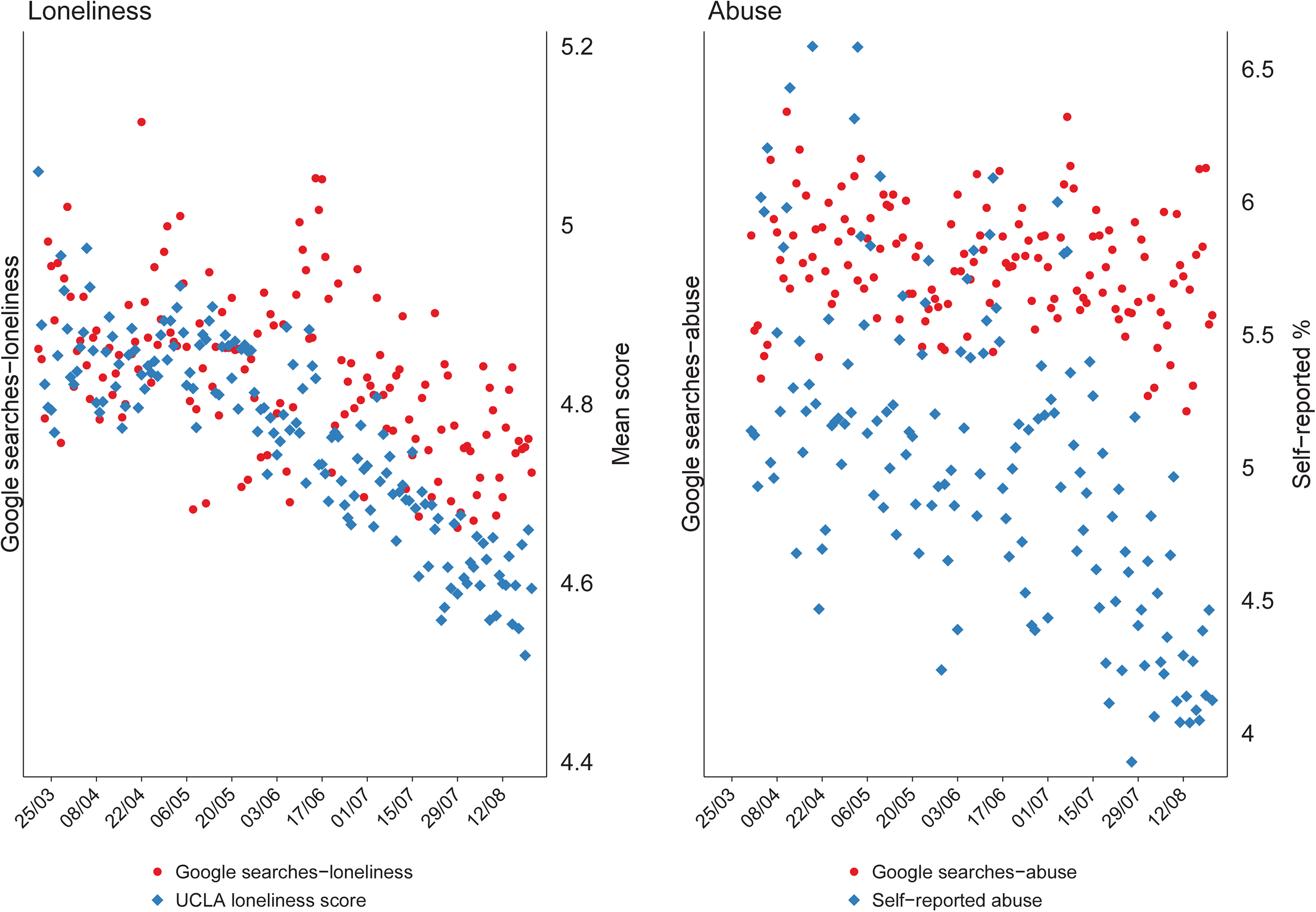
Self-reported and Google Trend search data for social distress topics in the UK

Table 1 presents the p-values of the Granger causality tests, which test whether the self-reported mental and social distress measures Granger-cause (i.e. predict) Google searching for the related search topic. For all but depression (p=0.03) and suicide (p=0.04), there was no evidence that changes in the self-reported mental distress measures were followed by changes in Google searching; Figure 1 indicates that these are inverse associations. There was some evidence that Google searches for the topic of suicidal ideation preceded self-reported self-harm (p=0.05), but Figure 1 indicates that this was an inverse association.

In relation to the measures of social distress, there was statistical and graphical evidence that the declines in reported levels (mean scores) of loneliness were associated with declines in Google searches (p<0.001).

When survey responses were stratified by age group, the declines in self-reported depression and anxiety appeared to be strongest in the younger age groups, with no clear age differences for the other mental and social distress markers (Figure 3 and 4).

**Figure 3.**
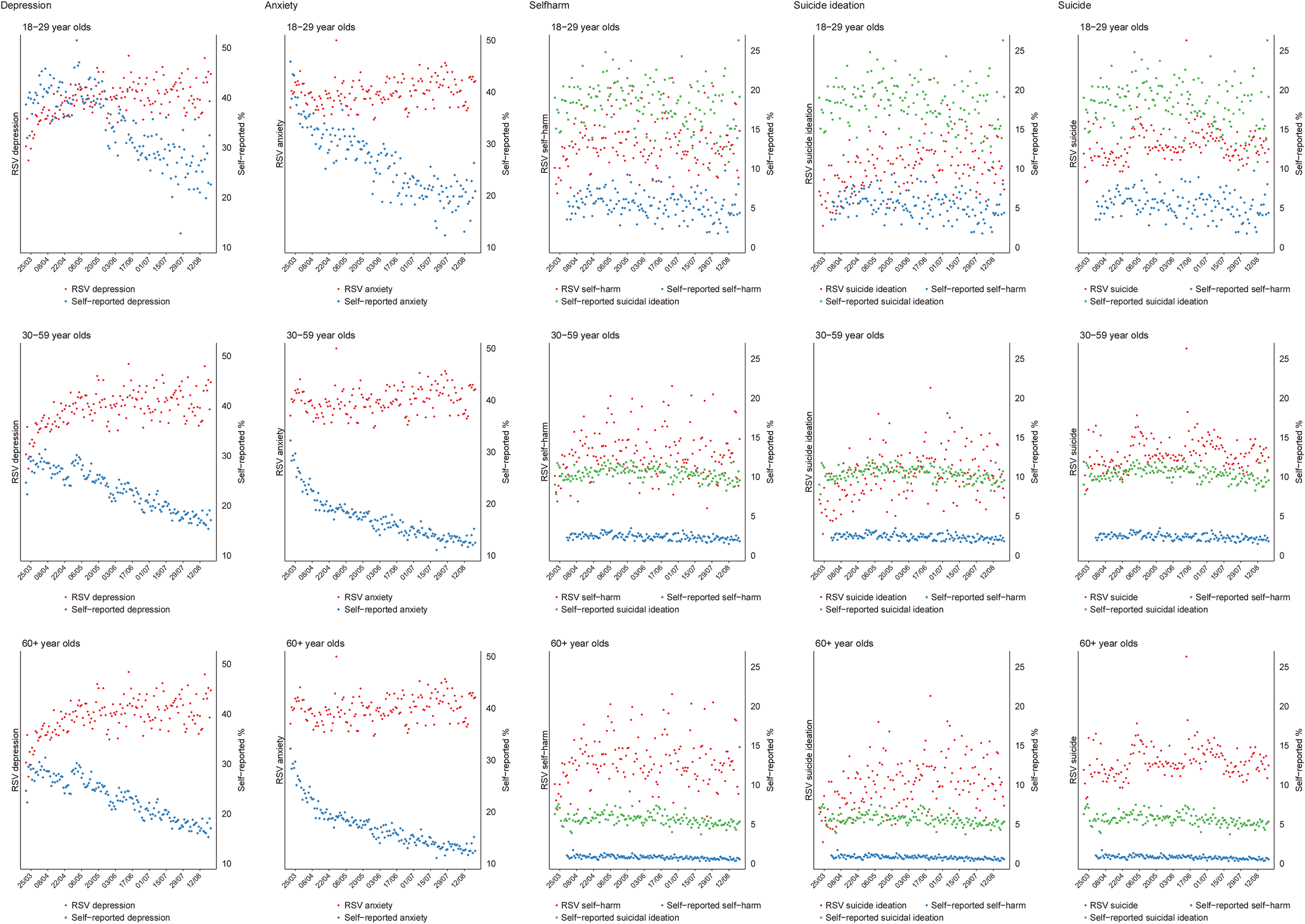
Self-reported and Google Trend* search data for mental distress topics in the UK stratified by age group *Google data are not available stratified by age and are presented here as overall values.

**Figure 4.**
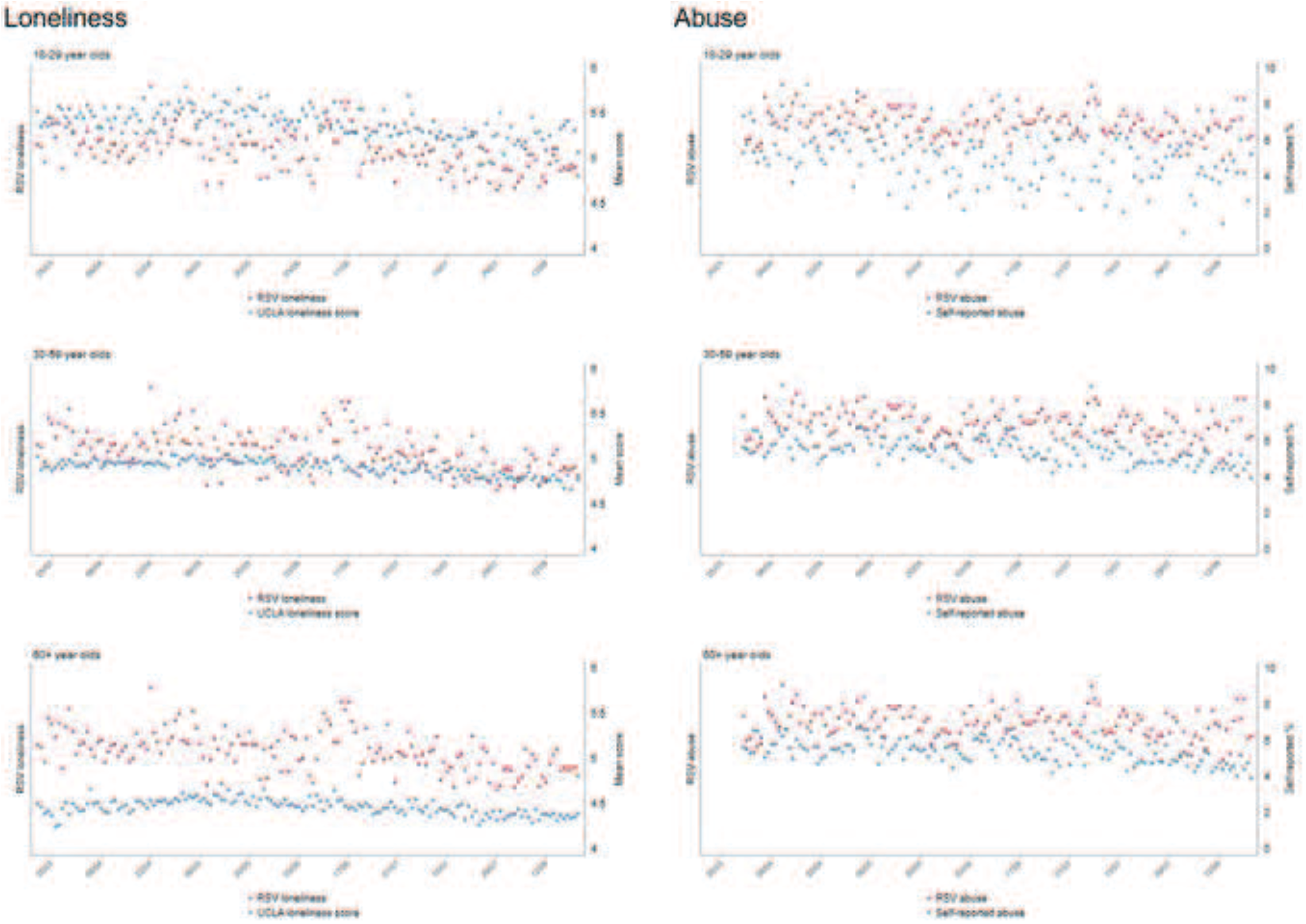
Self-reported and Google Trend* search data for social distress topics in the UK stratified by age group *Google data are not available stratified by age and are presented here as overall values.

## Discussion

Over the study period (21/03/2020-21/08/2020), population levels of self-reported depression and anxiety symptoms fell by around 40%, providing a powerful test of the responsiveness of Google Trends to changing population mental health. Nevertheless, we found no evidence of an association between self-reported anxiety and self-harm and Google Trends searching for these topics. There was some evidence of an association between self-reported depression and suicidal ideation with Google searches related to these topics, but this association was inverse: whilst trends in reported depression symptoms and suicidal ideation declined over the study period, relative search volumes for related topics in Google Trends increased. There was some evidence that Google searches for the topic of suicidal ideation preceded self-reported self-harm, but again, graphical evidence indicated that this was an inverse association. However, there was statistical and graphical evidence that Google searches for loneliness and self-reported mean scores for loneliness tracked one another.

The findings of a lack of a positive association between experiencing and searching on Google for terms relating to anxiety, depression, suicidal ideation and self-harm reflects concerns from some previous studies about using Google search terms as a tool to track population suicide rates(Page et al., 2011; Tran et al., 2017). Nevertheless, there is good evidence from person based studies that the Internet is used by suicidal individuals to search for methods of self-harm/suicide and also as a resource for help-seeking (Biddle et al., 2018; Padmanathan et al., 2018). There is also evidence of short-lived rises in relative search volumes for suicide-related terms following celebrity suicide deaths or suicide deaths using unusual methods, perhaps reflecting media interest rather than an increase in the number of suicidal individuals (Armstrong et al., 2021; Gunnell et al., 2015). However, there are several possible explanations for the generally null findings in our study. First, it could be that Google Trend data on topic-specific relative search volumes may be unreliable indictors of absolute search trends on those topics during a time with likely rapidly changing search volumes. Google do not provide data on absolute search volumes, and so it’s not possible to tell whether any changes in search trends are related to actual changes in volume of searches on specific topics, or due to changes in the denominator (i.e. the total number of Google searches on any day). It would be reasonable to hypothesise that during the pandemic (especially during the early stages), search volumes would have increased considerably due to a collective desire to learn more about the current situation, stay-at-home orders and working from home and this may explain the unexpected inverse associations seen between Google searches and reported symptoms of depression and suicidal ideation. Second, it could be that findings in relation to suicide deaths do not extend to symptoms of mental illness. The internet may be less used as a resource for individuals experiencing anxiety or depression during a crisis situation, especially as engaging with the media during this pandemic has been shown to predict worse levels of symptoms of depression and anxiety (Bu et al., 2020) Lastly, it is important to remember that Google Trends data have not been developed as a Public Health surveillance tool, and it could be that the components used by Google Trends for topics such as depression and anxiety are less specific to these mental health disorders than are those for suicide and some other topics.

Due to time delays in the publication of official suicide statistics we were unable to investigate associations between the incidence of suicide and Google searches on the topic of suicide. Data from real time surveillance of suicide trends in England for a population of approximately 9 million indicate that suicide trends were relatively stable up to August 2020 (National Confidential Inquiry into Suicide and Safety in Mental Health, 2020) – a finding which is broadly consistent with the Google Trends data.

It is notable that there was an association between self-reported loneliness and Google searches for loneliness, despite relative search volumes for loneliness being affected by the same factors (i.e. rises in total searching) that may have influenced the trends we saw for depression, anxiety and suicide-related outcomes (see above). This should be viewed with some caution in view of the multiple associations we investigated. There may be additional methodological points to consider when interpreting this finding. Firstly, the search volumes for the topic loneliness are likely to be lower than the other indicators. To get a sense of this we compared (post hoc) the Google Trends topic of loneliness against a benchmark/control topic (i.e., a topic with stable and predictable search volumes overtime – in this case the topic internet) (Carneiro and Mylonakis, 2009; Fowle, 2020). We observed that the RSVs for the loneliness topic to be considerably lower than the control topic, and the other mental and social distress topics (with the exception of self-harm and suicide ideation which showed similar levels). The possible low search volumes indicate unstable trends and should be interpreted with caution. Secondly, whilst the other mental and social distress topics were compared to the percentage of self-reports of their corresponding measures, the loneliness topic was compared to the mean score of the UCLA-3 questionnaire. There are no recommended cut-offs for identifying loneliness with this scale, and therefore the comparison may have low validity. If, however, the association is real, it is possible that searching for loneliness differs from searching in relation to symptoms of mental health conditions. A lonely person may, for example, use the Internet to make connections with others. More in-depth research to understand the sites identified and used by people searching on terms related to loneliness would help better understand this

### Strengths and Limitations

The availability of validated measures of population mental and social distress in a large population sample measured at weekly intervals, over a period when there were marked changes in levels of population distress and risk factors for poor mental health, gave us a powerful opportunity to identify whether changes in mental health were mirrored in Google search activity. It is unusual for population mental health to fluctuate so markedly over a short period of time, so the context of the pandemic provided an important natural experiment for testing the usefulness of Google search activity as a research tool within mental health. Nevertheless, it is possible that search activity and factors concerning individuals may be very different during a pandemic and associated public health measured than at other times.

There are several limitations to the analysis. First, the sociodemographic characteristics of survey responders and Google users differ. The COVID-19 Social Study is a sample of individuals who volunteered to complete weekly surveys. Young people, males, ethnic minorities, and people with low educational levels were relatively under-represented in the survey. In contrast the sociodemographic characteristics of Google users are unknown, but in the UK Internet use declines with age and is less frequent amongst those with disabilities (Office for National Statistics, 2019). Second, in our statistical models we compared survey responses on a specific day vs. Google Trends RSVs after a few days (i.e., lagged associations); however survey questions about mental and social distress ask about symptoms and events over the preceding week, so survey responses may not have reflected a participant’s feelings on the day they responded, and may not relate to future search behaviour. Third, self-harm is often carried out with no suicidal intent, and so questions about self-harm may be poor indicators of suicidal thoughts and behaviour. Fourth, our analysis is restricted to looking at trends following the onset of the pandemic. Extending the Google Trend data to include datapoints from January 2020 (as we have done previously (Knipe et al., 2020b)) indicates that after the first death in the UK the relative search volumes for depression topics markedly fell before returning to pre-pandemic levels. The rise in depression searches observed in this current analysis, therefore, represents a ‘bounce back’. Without objective mental health data prior to the pandemic period it is difficult to put the changes observed into context. It could be that levels of mental distress were continuing on an already established downward trajectory prior to the pandemic, or the declines observed could reflect levels returning to pre-pandemic levels. If either is the case, the Google Trends data for mental distress still does not track self-reported data. Fifth, the precise search terms contributing to the Google Trends topics are not specified, so it is possible that some searching related to these may have been missed; more detailed analyses focusing on specific topics and working with people with these conditions is warranted. Lastly, as a high proportion of our sample scored over 10 on the PHQ-9 (depression) and GAD-7 (anxiety) – 15% and 10% respectively - it is possible that we did not capture changing levels of those with much more severe symptoms. Higher symptoms levels / impairment are more likely to prompt help-seeking from health professionals and the Internet.

### Public Health Implications

Google Trend data, as currently formulated, do not appear to be a useful indicator of changing levels of population mental distress during a major public health crisis. It is likely that a range of other factors – such as the total volume of searching, news reporting stimulating curiosity about mental health topics and the impact of the development of mental health symptoms on engagement with the Internet - may influence relative search volumes and activity for these terms. It is too early to judge the utility of Google trends as an indicator of suicide rates as previous studies have shown mixed results and we found no positive associations with indicators of suicidal behaviour. Indeed, there was some evidence of an inverse association between self-reported suicidal ideation and Google searching for suicide and self-harm, suggesting that we should be especially wary about using Google search terms as tools for monitoring self-harm within the population. However, the positive association we found with loneliness should be explored further. Overall, the findings of this study urge caution when attempting to utilise Google Trends data as a public health surveillance tool for tracking population mental health.

## Supporting information

Graphical abstract

## Data Availability

Data on Google Trends are publicly available. UCL COVID Social Study data will be made publicly available following the end of the pandemic.

