## Supplementary material for "Is Google Trends a useful tool for tracking mental and social distress during a public health emergency? A time-series analysis": Graphical abstract

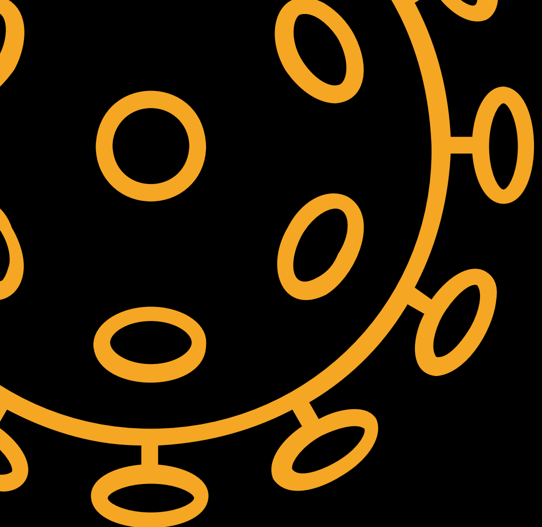

Can Google searches be used as a mental health tracking tool?

Using survey data from 70,000+ people in the UK we validated data from Google Trends

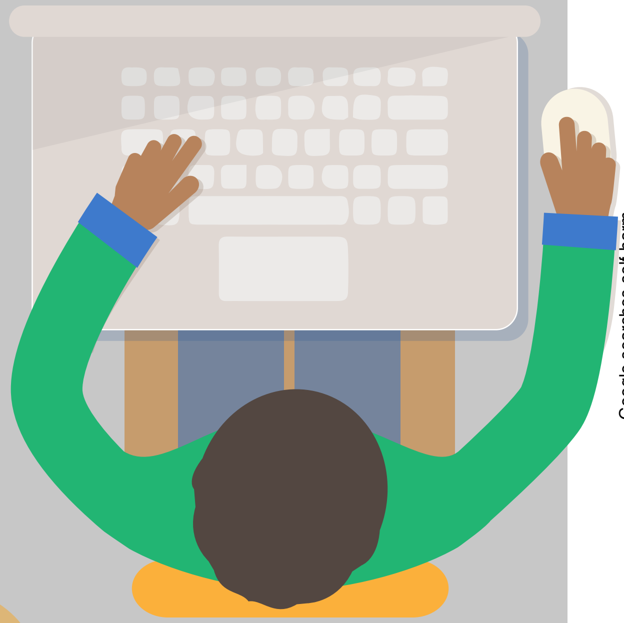

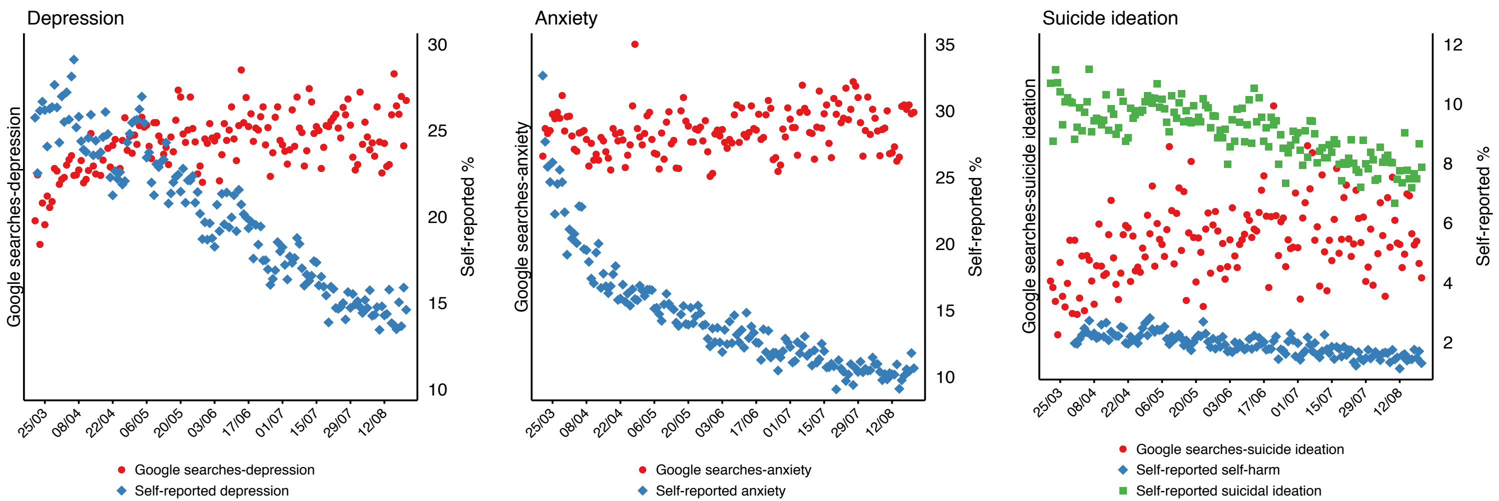


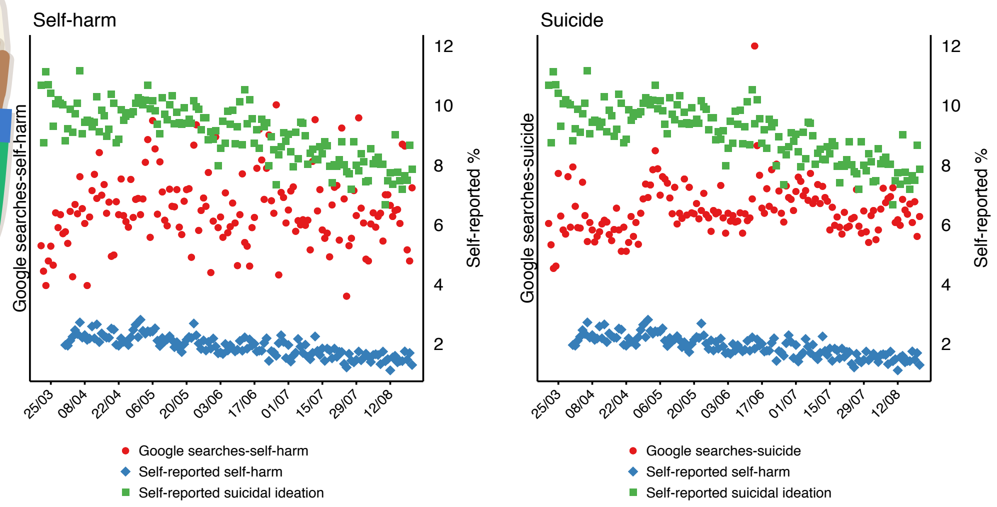


Google Trends did not track measured mental health during the COVID-19 pandemic
